# Competing endogenous RNA networks reveal long non-coding RNAs as potential prognostic biomarkers in gastric cancer: A systematic review and meta-analysis

**DOI:** 10.1101/2023.04.22.23288939

**Authors:** Sadra Salehi-Mazandarani, Mohammad Hossein Donyavi, Amirhossein Vedaei, Alireza Najimi, Ziba Farajzadegan, Parvaneh Nikpour

## Abstract

**Aims:** Gastric cancer (GC) represents a global health-care challenge. In recent years, a large number of competing endogenous RNA (ceRNA) network studies have elucidated critical long non-coding RNAs (lncRNAs) as potential prognostic biomarkers in GC. Nevertheless, there is no systematic review and meta-analysis regarding them. Here, we propose a systematic review and meta-analysis for lncRNAs as prognostic biomarkers based on the ceRNA network studies in GC.

**Main methods:** The Web of Science, Embase, PubMed, Scopus, ProQuest and Google Scholar databases were searched to collect eligible ceRNA network studies in which lncRNAs were reported as prognostic biomarkers until 27^th^ June 2022. The pooled hazard ratios (HRs) were assessed to evaluate the prognostic effect of the lncRNAs. Prognostic values of highly reported lncRNAs were also examined. Furthermore, sensitivity analysis, and publication bias were evaluated.

**Key findings:** Totally, 35 studies were included in the systematic review among which 13 studies were eligible for meta-analysis. Pooled examination of HRs based on the univariate and multivariate reports revealed that the expression changes of the lncRNAs correlate with overall survival (OS) of GC patients. Two lncRNAs including *CCDC144NL-AS1* and *LINC01094* were highly reported as potential prognostic biomarkers based on the ceRNA network studies. Prognostic value of these two lncRNAs was furthermore confirmed by meta-analysis.

**Significance:** Our meta-analysis demonstrated potential application of lncRNAs introduced based on the ceRNA network studies as promising prognostic biomarkers in GC. Further functional studies are needed to elucidate the molecular mechanisms underlying these lncRNAs’ functions in GC.

## 1. Introduction

According to the information of the Global Cancer Observatory in 2020, gastric cancer (GC) had the sixth and fifth rates of incidence and mortality among the different types of cancers, respectively (1). Furthermore, GC was the sixth most prevalent cancer worldwide (1). The incidence of GC is variable among different countries and cultures (2). Most of the cases are diagnosed in eastern countries such as Japan and Korea where the population screening of GC is conducting. Although earlier detection and reduced *helicobacter pylori* (*HP*) infection in these countries have led to the decreased incidence of GC, it is still a serious concern (3). The 5-year survival rate of advanced GC in western countries, in which GC is mostly diagnosed at an advanced stage, is less than 10% (4). GC is a complex disease and many factors participate in its initiation and progression. The risk of GC has a direct correlation with increased ages and is higher in men rather than women. *HP* infects nearly half of the world□s population and is a primary risk factor in GC. Some other risk factors for this type of cancer include blood type A, familial adenomatous polyposis (FAP) syndrome, Peutz–Jegher syndrome (PJS), salt intake, smoking, and alcohol (5).

Identification of novel non-invasive prognostic biomarkers with high sensitivity and specificity has always been a hot topic in the field of cancer research. Therefore, many studies have introduced novel prognostic biomarkers and evaluating their accuracy in GC. One of the promising type of prognostic biomarkers in GC which have recently received great attention are long non-coding RNAs (lncRNAs) (6).

lncRNAs are a group of non-coding RNAs with the length of longer than 200 nucleotides (7). Although they are not translated to proteins, they have critical physiological roles in cellular differentiation, stress, aging, proliferation, apoptosis and transcriptional regulation (8). Dysregulation of lncRNAs in cancers including GC have been identified (6). They affect tumor formation, progression, and metastasis (9). One of the most studied ways by which lncRNAs participate in tumorigenesis is their role as competing endogenous RNAs (ceRNAs). In fact, lncRNAs as well as other types of RNAs such as messenger RNAs (mRNAs) and circular RNAs (circRNA) which have the same microRNA (miRNA) response elements (MREs) compete with each other for binding to the miRNAs (10). These crosstalk among various kinds of RNAs form the ceRNA networks that can promote tumorigenesis. The study of these networks can provide a deep view of molecular and cellular characteristics underlying GC progression and introduce novel prognostic biomarkers.

Over the last years, many studies have been conducted to identify ceRNA networks in GC. These studies mostly have some special steps consisting of data retrieval, identification of differentially-expressed RNAs (DERs), prediction of DERs interactions, ceRNA network construction, functional enrichment and finally survival analyses. Because of high accessibility to the different methods and databases for survival analysis of the RNA expression data, survival analysis has been somehow an inseparable part of ceRNA network studies. Utilizing The Cancer Genome Atlas (TCGA) (11) and gene expression omnibus (GEO) (12) data at most of the times, which have large sample sizes, can make the results of these studies, highly reliable. The survival analyses in these types of studies, have revealed the correlation between many lncRNAs and overall survival (OS) of GC patients. Due to the lack of a systematic review and meta-analysis regarding lncRNAs which have been reported as potential prognostic biomarkers from ceRNA network studies in GC, this meta-analysis set out to investigate the prognostic value of these lncRNAs in GC patients.

## 2. Materials and methods

### 2.1. Guideline and registration

This study was conducted based on the Preferred Reporting Items for Systematic Reviews and Meta-Analyses statement (PRISMA) (13). The review protocol was registered on The International Prospective Register of Systematic Reviews (PROSPERO) (CRD42022360864).

### 2.2. Review Question

Which lncRNAs have been introduced as potential prognostic biomarkers in GC according to ceRNA network studies, and how much is their prognostic effect?

### 2.3. Selection criteria

Inclusion criteria included:

1. Having a ceRNA network including lncRNAs in GC
2. The number of at least one type of RNAs (lncRNA, miRNA, mRNA, etc) in the ceRNA network should be more than 1.
3. OS has been assessed.
4. Studies based on human samples

Exclusion criteria included:

1. Review studies
2. Studies not in English
3. Retracted studies
4. Studies not specified to GC
5. Studies without univariate survival analysis or its detailed results (after correspondence)
6. Congresses abstracts
7. Studies regarding a specific state of GC (For instance examination of *HP* infection in GC)

### 2.4. Literature Search Strategy

Online literature search was conducted up to 27^th^ June 2022 in the Web of Science, Embase, PubMed, Scopus and Cochrane Central Library using a developed search query (Supplementary file 1). Grey literature was also searched on ProQuest and Google Scholar. Furthermore, reference lists of included studies were manually searched for studies potentially meeting the eligibility criteria. In case of missing information, authors were contacted via email.

### 2.5. Screening, quality assessment and data extraction

Records were imported to EndNote software ver. X8.0.1 and duplicate studies were removed. Two reviewers (SSM and MHD) independently screened title and abstracts of the studies to select relevant studies. Then, the full texts of the selected studies were assessed against inclusion and exclusion criteria. Any disagreement was solved through discussion with another reviewer (PN) until a consensus was reached.

After full text examination, the quality of the remained studies was assessed by Joanna Briggs Institute (JBI) critical appraisal checklist for cohort studies (14). Studies reached score of > 7 (each “yes” answer was considered as one score and “no” answers as zero), were selected as high quality and entered to the next step of data extraction.

An excel datasheet was designed for data extraction and two reviewers (SSM and MHD) independently extracted the data from the included studies. Extracted information consist of the name of the first author, year of publication, country of origin (the country of the corresponding author), origin of samples, sample size, the name of lncRNA(s), hazard ratio (HR), *p*-value, 95% confidence interval (CI), follow-up time and references. A third reviewer (PN) was consulted in case of disagreement. Univariate as well as multivariate OS analyses reports in which the correlation of one lncRNA with confounding factors such as age, gender, stage, etc were evaluated, were considered in the current study. Basis of HR examination (high expression group of lncRNA divided by low group or vice versa) were also considered.

### 2.6. Statistical analysis

All analyses were performed using STATA software ver. 11.0. In all of the included studies to the meta-analysis part, the basis of HR examination was clear and similar (high expression group/low expression group). Two studies with unclear basis of HR calculation and the reference group (even after mailing) were excluded in meta-analysis (15, 16). The pooled examination of HRs with 95% CI of univariate survival results was performed based on the 11 studies, in which sample sizes were reported (17-27). Besides, according to the information of multivariate survival analysis, the pooled HRs with 95% CI was calculated. Four studies (23, 27-29) with clear basis of HR examination were considered for pooled examination of multivariate reports. Furthermore, lncRNAs which were reported at least three times according to the univariate or multivariate survival analysis were meta-analized.

Forest plots were utilized to estimate the correlation of these lncRNAs expression with OS of GC patients.

*I*^2^ index was used to assess the between-study heterogeneity, If the heterogeneity was substantial (*I*^2^>50%, *p*-value<0.05), the random effect model would be selected, otherwise, the fixed effect model would be utilized. lncRNAs with HR>1 and HR<1 were interpreted as risk and protective factors, respectively. For lncRNAs as risk factors, GC patients with higher expression of them, have shorter OS while for lncRNAs as protective factors, GC patients with higher expression of them have longer OS. *P*-value<0.05 was considered as statistically significant for the survival results. In addition, Begg’s test was performed to assess the publication bias (30) and *p*-value<0.05 were considered as significant publication bias.

Sensitivity analysis was performed for the lncRNAs obtained from multivariate analyses as well as lncRNAs which had been studied at least three times including univariate and multivariate analyses.

## 3. Results

### 3.1. Characteristics of the included studies

Out of 1764 initially identified studies through online and manual search, 1019 duplicates were removed, 684 studies were removed when screening title and abstracts and 21 studies were excluded though full-text reading. The quality of 40 remaining studies were examined and 5 studies did not pass this step due to having low quality (quality score ≤7) (Supplementary file 2). Overall, 35 studies were recruited in the current study after selection by the mentioned criteria. The number of records at each step of the study selection process is shown in PRISMA flow chart (Figure 1).

**Figure 1.**
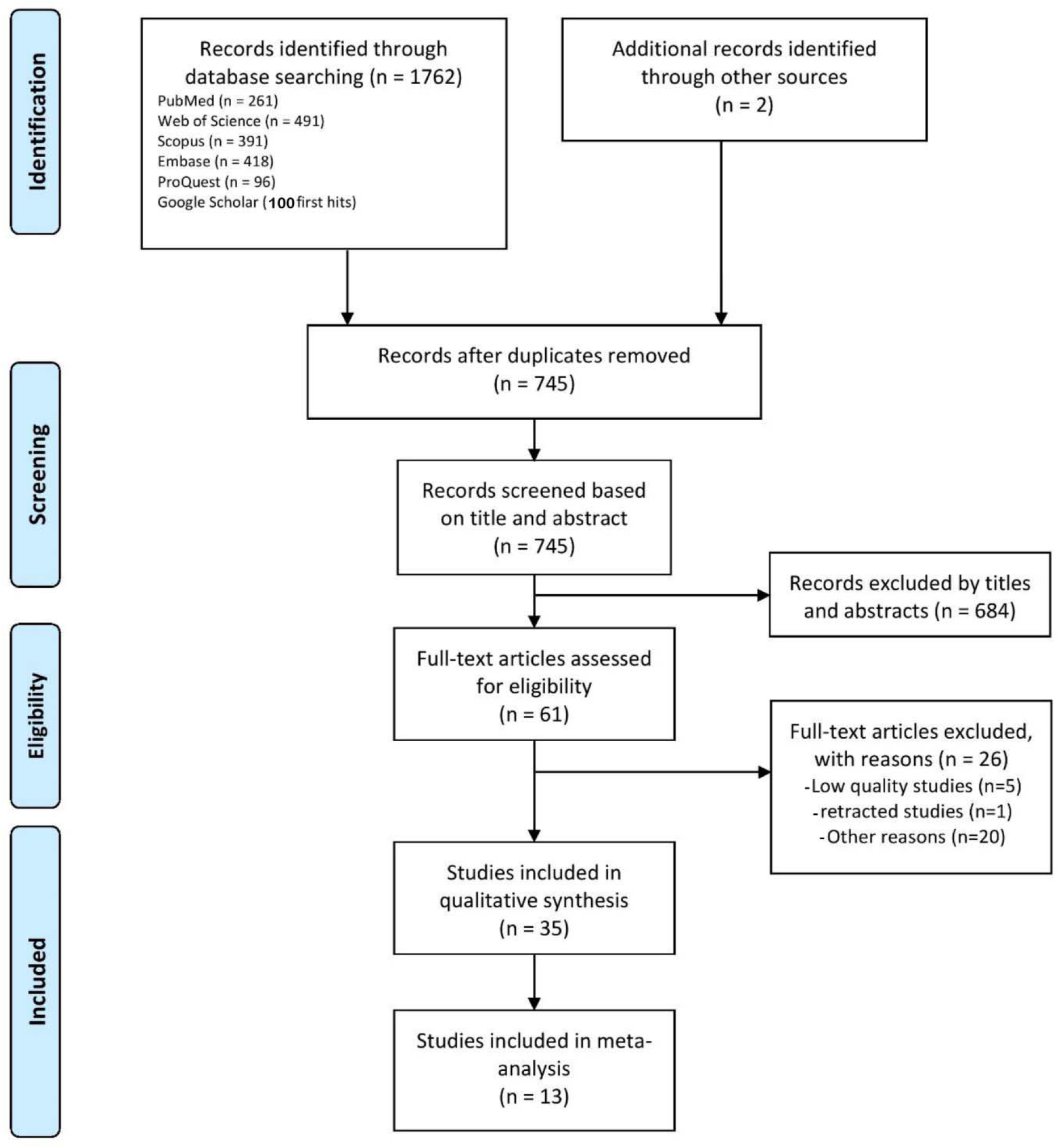
The Preferred Reporting Items for Systematic Reviews and Meta-Analyses (PRISMA) flow diagram of the search results and number of records at each stage

Excluded studies with the reason of exclusion are reported in supplementary file 3. Except one study (31), others had reported statistically significant prognostic lncRNAs (*p*-value<0.05), based on univariate survival analyses, which have been presented in table 1.

**Table 1.**
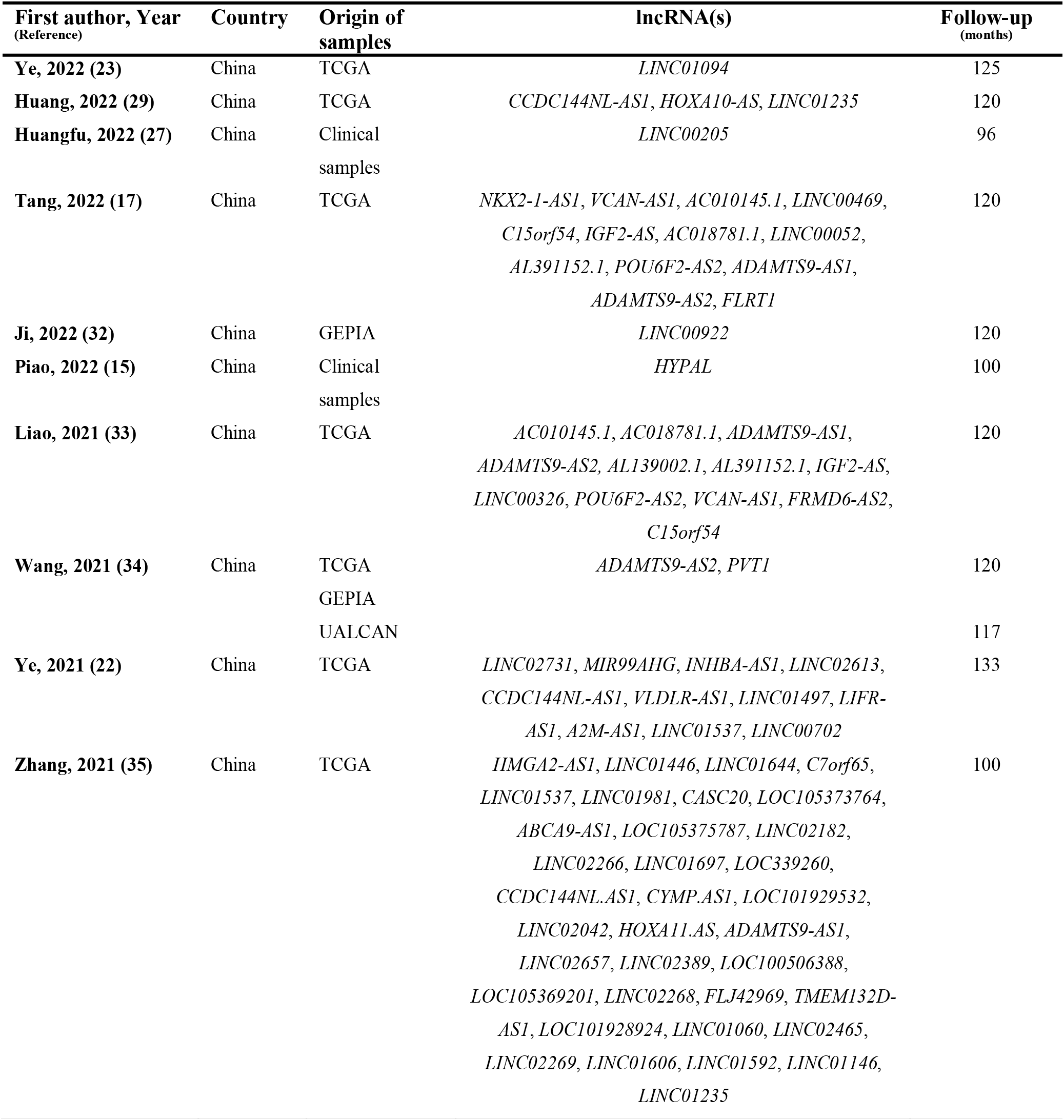
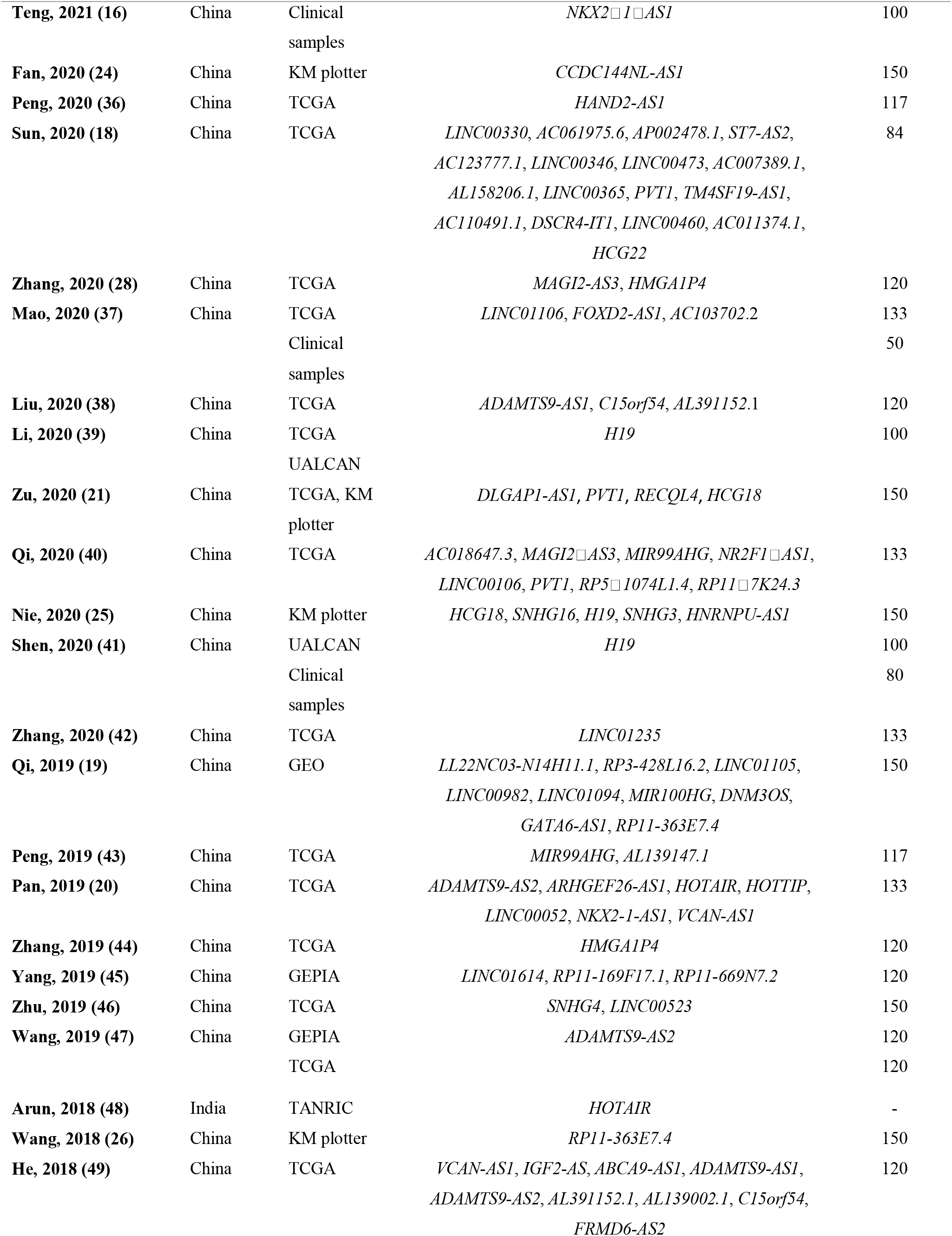
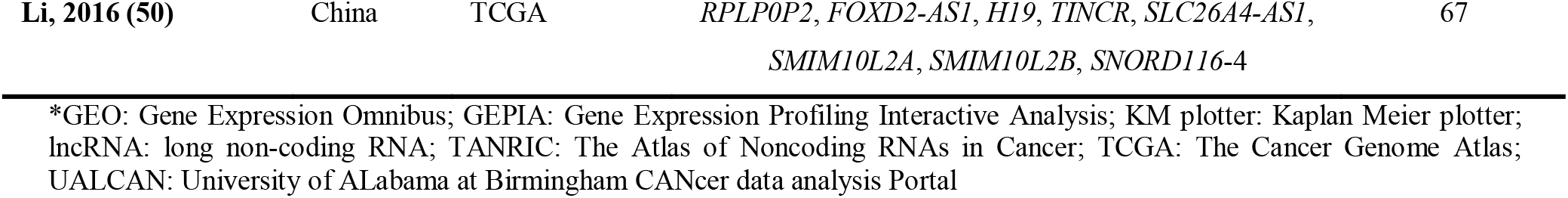
Significant prognostic value of the lncRNAs mentioned in the ceRNA network studies based on univariate survival analyses

Supplementary file 4 contains the main characteristics of the 35 included studies. In all of the studies, a ceRNA network including lncRNAs were constructed and univariate/multivariate survival analysis were performed to identify correlation of lncRNA(s) with OS in GC. In table 1 and supplementary file 4, HR>1 and HR<1 represent shorter OS in patients with higher and lower expression of lncRNAs, respectively (The basis of HR examination in two studies (15, 16) were not clear). Six studies performed multivariate survival analysis based on evaluating the effect of one lncRNA with cofounding factors (Table 2).

**Table 2.** Results of multivariate analysis in which the prognostic effect of one lncRNA has been analyzed with confounding factors

| Signature | HR | P-value | CI | Ref. |
| --- | --- | --- | --- | --- |
| **T stage** | 0.732 | 0.305 | 0.404-1.329 | (23) |
| **N stage** | 0.865 | 0.672 | 0.443-1.690 |  |
| **M stage** | 0.868 | 0.734 | 0.384-1.962 |  |
| **Histologic grade** | 0.663 | 0.068 | 0.426-1.031 |  |
| **Pathologic grade** | 0.788 | 0.437 | 0.432-1.437 |  |
| **Age** | 0.591 | 0.013 | 0.391-0.893 |  |
| **Primary therapy outcome** | 4.313 | <.001 | 2.867-6.489 |  |
| ***LINC01094*** | 1.535 | 0.039 | 1.021-2.308 |  |
| **Age** | 1.82 | 0.001 | 1.26-2.63 | (29) |
| **Pathologic stage** | 1.32 | 0.333 | 0.75-2.34 |  |
| **T stage** | 1.27 | 0.372 | 0.75-2.13 |  |
| **N stage** | 1.37 | 0.190 | 0.86-2.2 |  |
| **M stage** | 2.32 | 0.008 | 1.24-4.32 |  |
| ***CCDC144NL-AS1*** | 1.79 | 0.001 | 1.25-2.57 |  |
| **Vascular invasion** | 2.215 | 0.038 | 1.043-4.703 | (27) |
| **histological differentiation** | 0.528 | 0.175 | 0.210-1.329 |  |
| **tumor size** | 0.956 | 0.892 | 0.497-1.837 |  |
| **age** | 0.988 | 0.970 | 0.520-1.879 |  |
| **T** | 0.923 | 0.858 | 0.385-2.212 |  |
| **N** | 0.763 | 0.669 | 0.220-2.644 |  |
| **M** | 6.674 | 0.006 | 1.713-26.001 |  |
| **stage** | 3.797 | 0.008 | 1.413-10.204 |  |
| *Linc00205* | 3.273 | <0.001 | 1.652-6.592 |  |
| **TNM stage** | 2.745 | 0.004 | 1.575-5.136 | (16) |
| **Distant metastasis** | 2.054 | 0.008 | 1.201-3.634 |  |
| **Infiltration of peritumoral tissues** | 1.934 | 0.003 | 1.245-3.121 |  |
| *NKX2-1-AS1* | 0.383 | 0.005 | 0.297-0.501 |  |
| **Stage** | 1.429 | 0.102 | 0.931-2.194 | (28) |
| **Topography** | 0.973 | 0.865 | 0.710-1.334 |  |
| **Metastasis** | 1.317 | 0.502 | 0.589-2.941 |  |
| **Lymph node stage** | 1.084 | 0.518 | 0.850-1.382 |  |
| *MAGI2-AS3* | 1.472 | 0.023 | 1.055-2.055 |  |
| **Bormann** | 1.14 | 0.60 | 0.70-1.84 | (15) |
| **Tumor size** | 0.83 | 0.64 | 0.39-1.79 |  |
| **TNM stage** | 3.65 | 0.00 | 1.82-7.30 |  |
| *HYPAL* | 0.27 | 0.00 | 0.12-0.59 |  |

\*CI: confidence intervals; HR: hazard ratios; lncRNA: long non-coding RNA; M stage: Distant metastasis; N stage: Regional lymph nodes; T stage: primary tumor; TNM: Tumor, Node, and Metastasis

The original data of most of the studies were retrieved from TCGA database. The average follow-up time of the studies was 120 months. Except one (48), other studies were conducted in China. All articles were published between 2016 and 2022. The sample sizes among the included studies ranged from 50 to 876. In overall, studies reported statistically significant correlation of 122 unique lncRNAs with the prognosis of GC patients. Some of the studies used clinical samples and others utilized cancer databases including TCGA, GEO, GEPIA (51), UALCAN (52), KM plotter (53), and TANRIC (54).

### 3.2. The prognostic significance of lncRNAs

Among studies with univariate survival analysis, 11 studies with reported sample size, HR (with clear basis of calculation), and CI were identified for meta-analysis (17-27). These studies included 70 lncRNAs (including unique and repeated lncRNAs). Higher expression of 57 and lower expression of 13 lncRNAs were associated with shorter OS. Studies with multivariate analysis reported four lncRNAs with clear basis of HR including *LINC01094, CCDC144NL-AS1, LINC00205*, and *MAGI2-AS3* (23, 27-29). The pooled HRs for univariate reports was 1.01 (95% CI 1.01-1.02, *p*-value<0.001, Figure 2).

**Figure 2.**
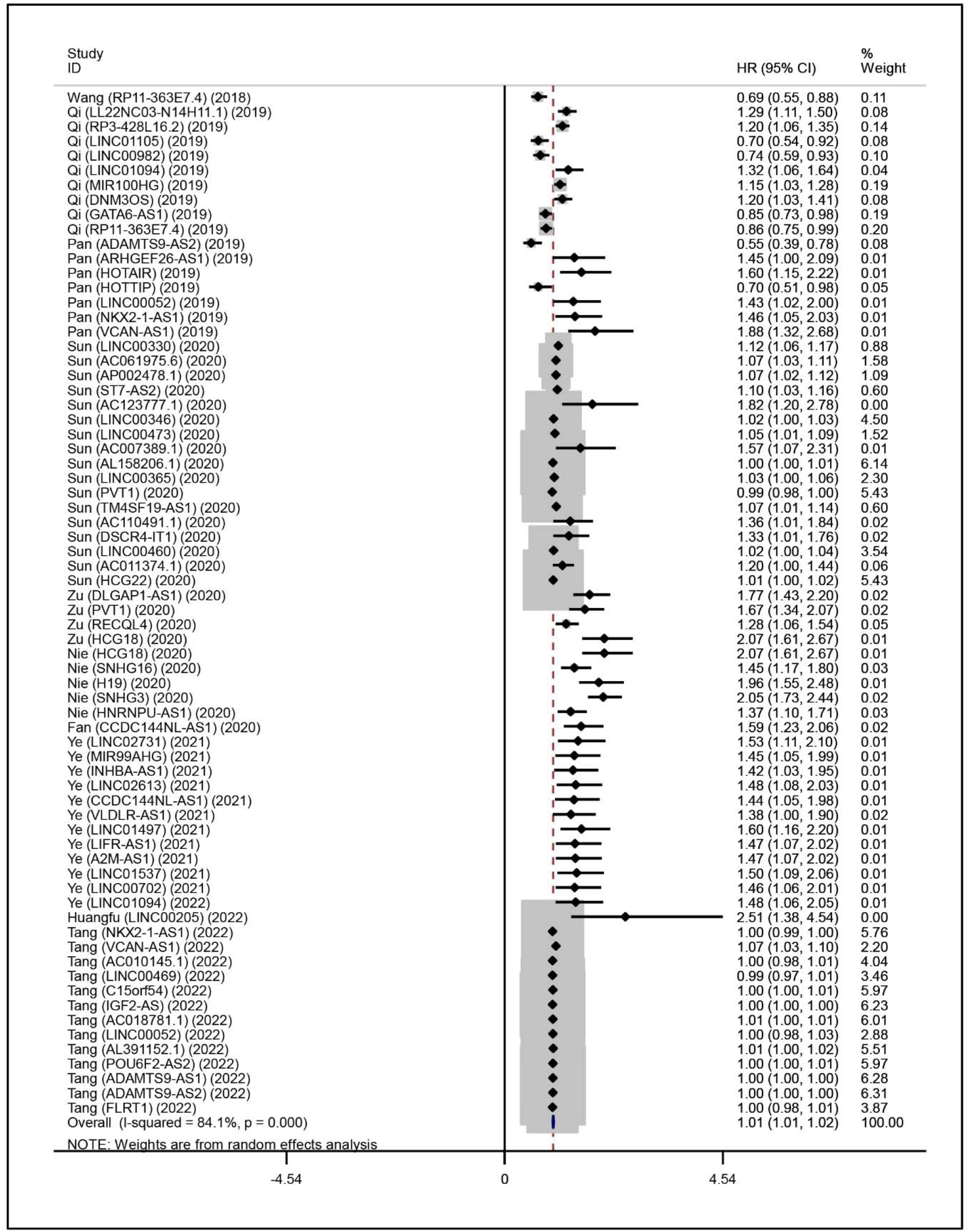
Forest plot of pooled examination of univariate survival results. CI: confidence interval; HR: hazard ratio

The pooled HRs for multivariate was 1.60 (95% CI 1.27-1.94, *p*-value<0.001, Figure 3A).

**Figure 3.**
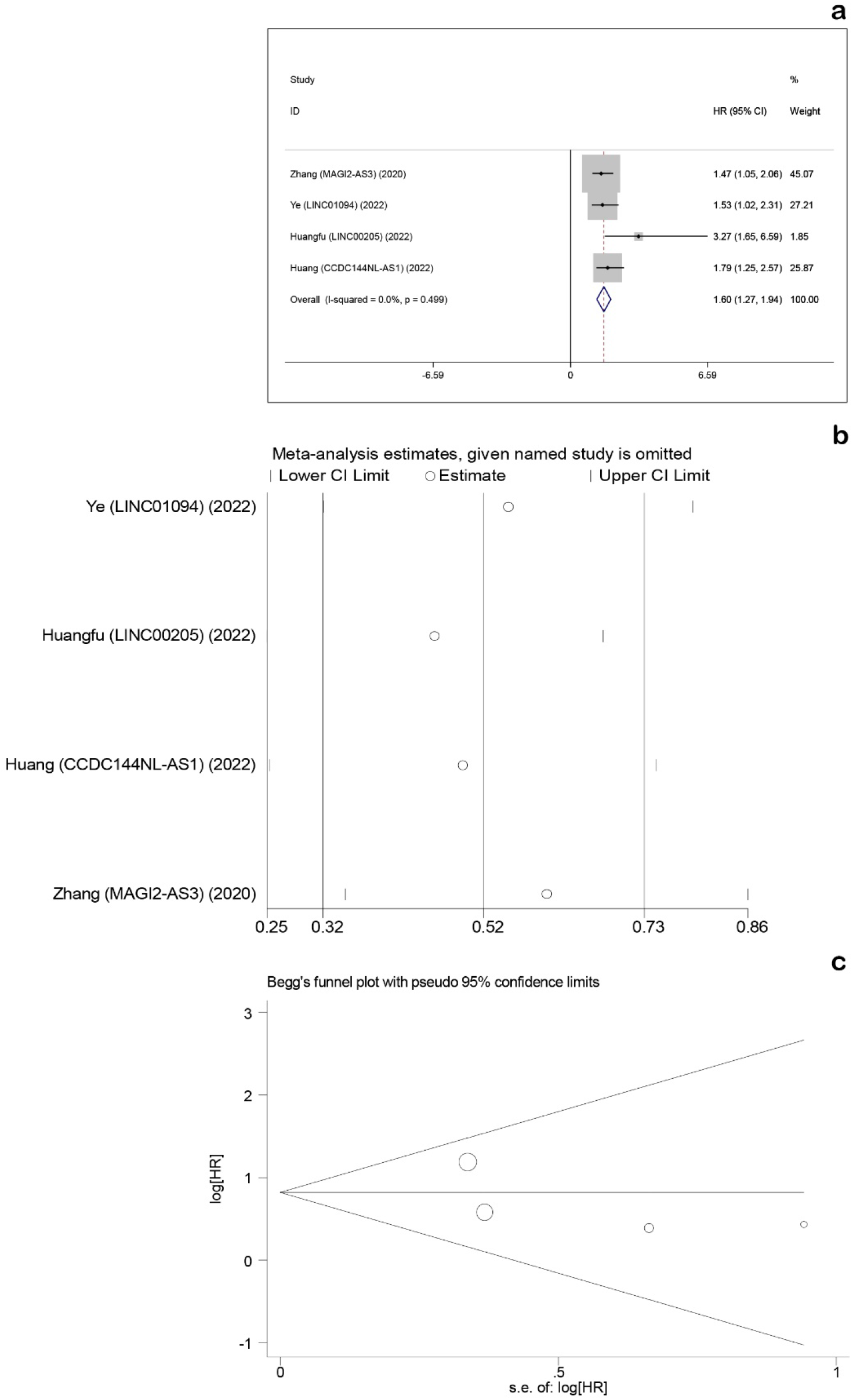
Forest, sensitivity, and funnel plots of multivariate studies, Pooled HRs were calculated for four studies with multivariate analysis (a), Sensitivity analysis was performed for four studies with multivariate analysis (b), Begg’s test was performed for four studies with multivariate analysis (c). HR: hazard ratio, lncRNA: long non-coding RNA CI: confidence interval

These results indicate that expression changes of the lncRNAs correlate with OS in GC and patients with higher expression of them have poorer survival.

### 3.3. *CCDC144NL-AS1* and *LINC01094* lncRNAs as prognostic biomarkers in GC

Two lncRNAs including *CCDC144NL-AS1* and *LINC01094* which were reported at least three times (based on the univariate and multivariate survival analysis) were identified. Forest plots of these lncRNAs have been presented (Figure 4a and 4b).

**Figure 4.**
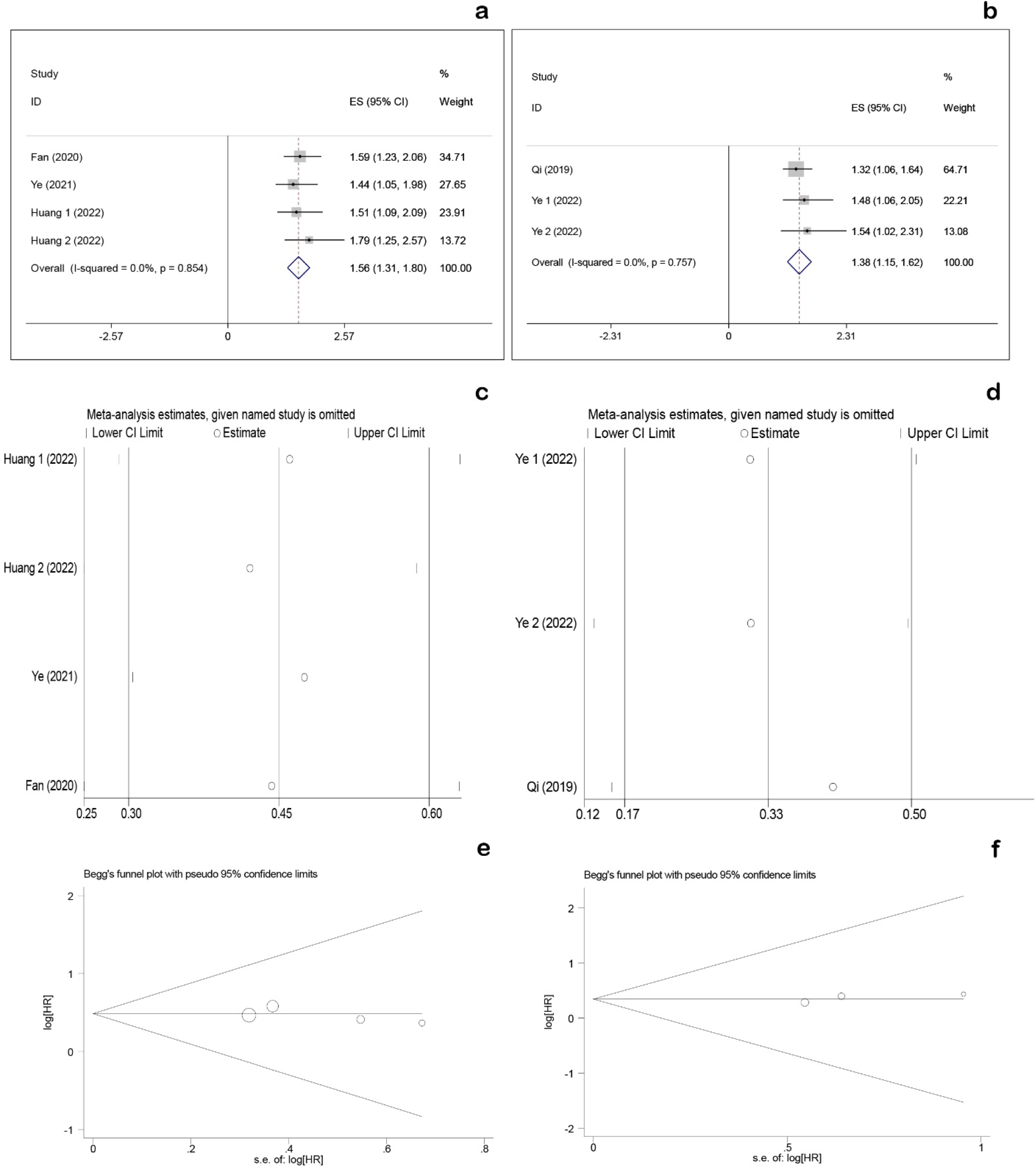
Forest, sensitivity, and funnel plots of two lncRNAs, Pooled HRs were calculated for *CCDC144NL-AS1* (a) and *LINC01094* (b), Sensitivity analysis was performed for *CCDC144NL-AS1* (c) and *LINC01094* (d), Begg’s test was performed for *CCDC144NL-AS1* (e) and *LINC01094* (f). HR: hazard ratio, lncRNA: long non-coding RNA CI: confidence interval

Three ceRNA network studies have reported prognostic effects of *CCDC144NL-AS1* based on three univariate and one multivariate analyses (22, 24, 29). On the other hand, *LINC01094* was reported according to two studies based on two univariate and one multivariate analyses (19, 23). Forest plots revealed that the expression of both *CCDC144NL-AS1* (HR=1.56, 95% CI: 1.31-1.80, *p*-value<0.001, Figure 3B) and *LINC01094* (HR = 1.38, 95% CI: 1.15-1.62, *p*-value<0.001, Figure 3C) have negative association with OS.

### 3.4. Sensitivity analysis

We performed sensitivity analysis for studies with multivariate analysis (Figure 3b) as well as those studies reporting *CCDC144NL-AS1* (Figure 4c) and *LINC01094* (Figure 4d) lncRNAs, respectively, even though no heterogeneity was detected (*I*^2^=0.0%, *p*-value>0.05). The results showed that this meta-analysis was reliable.

### 3.5. Publication bias

No significant publication bias was found in any of the univariate (Figure 5), multivariate (Figure 3c), and the two lncRNAs analyses (Figure 4e and 4f).

**Figure 5.**
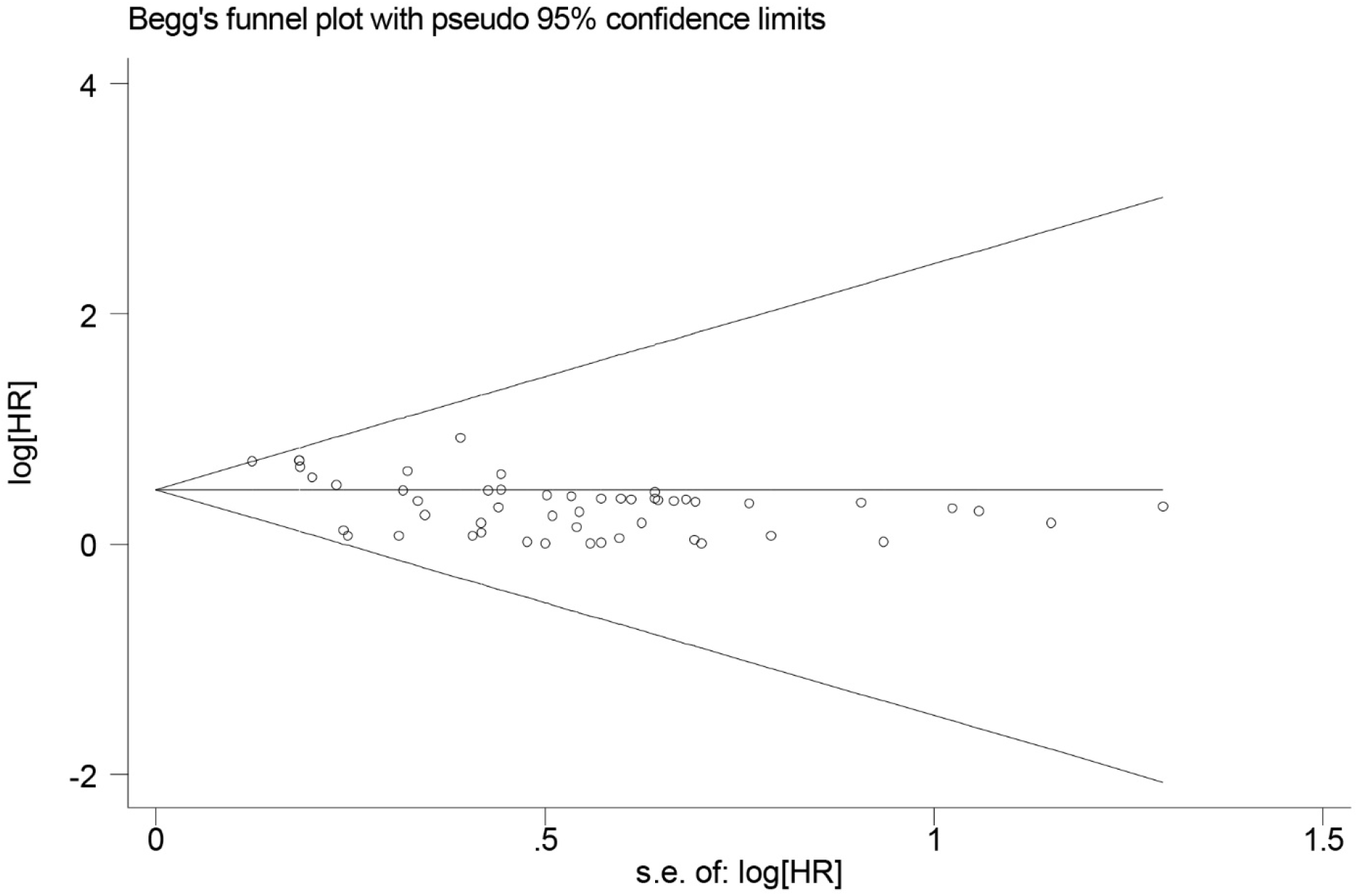
Funnel plot of pooled examination of studies with univariate survival results. HR: hazard ratio

The funnel plots are all symmetrical, demonstrating that there is no significant publication bias in this study. The *p*-value for Begg’s test of univariate, multivariate, *CCDC144NL-AS1* and *LINC01094* were 0.183, 0.923, 0.174 and 0.117, respectively.

## 4. Discussion

GC is one of the most common types of cancer around the world (1). Despite of currently treatment options such as surgery, chemotherapy, and radiotherapy, GC mainly reduces the quality of life (55). The overall relative survival rate of GC patients has been improved in the recent years, however it is still poor (56, 57). Therefore, identification of novel biomarkers to predict the survival rates of GC patients are needed.

In some ceRNA network studies, in which the competition among various types of RNAs for binding to the same miRNAs are examined, correlation of expression of ceRNAs with survival of GC patients has been evaluated (18, 21). To select the most relevant prognostic ceRNAs, researchers apply different strategies including selection of hub RNAs (mostly based on degree of centrality), differentially-expressed RNAs or RNAs interacting with other well-known RNAs.

Furthermore, most of these studies use databases like TCGA and GEO for data retrieval or analysis. These databases have mostly large sample size with long follow-up times which make their results highly reliable (58, 59). Up to now, a large number of ceRNA network studies have been published and many types of RNAs have been reported as potential prognostic biomarkers in GC. One of the most reported of these RNAs are lncRNAs. lncRNAs have been shown to be dysregulated in GC affecting GC cells progression, migration, metastasis, and apoptosis (60, 61).

To validate the accuracy of the reported lncRNAs as prognostic biomarkers in GC based on the ceRNA network studies, we conducted the current systematic review and meta-analysis. We searched literature looking for ceRNA network studies in GC which had reported lncRNAs as prognostic biomarkers. Thirty-five studies were with average follow-up time of approximately 120 months. Pooled examination of HRs based on both univariate and multivariate reports revealed that expression changes of lncRNAs, as ceRNAs, are correlated with OS of GC patients. Two lncRNAs entitled *CCDC144NL-AS1* and *LINC01094* were recurrently reported as GC prognostic biomarkers. Accordingly, we assessed prognostic values of them utilizing a meta-analysis approach and identified that overexpression of them correlates with poor OS in GC patients.

Among the included studies in the current meta-analysis, three (22, 24, 29) and two (19, 23) studies introduced *CCDC144NL-AS1* and *LINC01094* as prognostic biomarkers in GC, respectively. As mentioned above, three ceRNA network studies reported *CCDC144NL-AS1* as prognostic biomarker in GC. In 2022, Huang et al performed a ceRNA network study in GC and according to univariate and multivariate survival analysis, they introduced *CCDC144NL-AS1* as a potential prognostic biomarker as its high expression was significantly associated with short OS (HR=1.5 (CI:1.09-2.09), Log-rank P=0.014) (29). Ye et al examined 378 GC samples from TCGA data and discovered *CCDC144NL-AS1* to be a potential biomarker for predicting GC prognosis. This was confirmed by univariate survival analysis (HR=1.4 (CI:1.04-1.99), *p*-value=0.018) and multivariate cox analysis (HR=1.4 (CI:1.04-1.99), *p*-value=0.006). Furthermore, this study discovered that this lncRNA affects proliferation, migration, and invasion of GC cells (22). In another study, researchers found that the expression level of *CCDC144NL-AS1* is higher in GC compared to normal tissues. This increase in expression is linked to the size of the tumor, a high rate of lymphatic spread, and a lower overall survival with a hazard ratio of 1.6 (CI:1.23-2.06) and a log-rank P value of 0.0035. Moreover, overexpression of *CCDC144NL-AS1* stimulates cellular proliferation, invasion, and migration of cancer cells in GC (24).

On the other hand, *LINC01094* was reported in two ceRNA network studies as a promising prognostic biomarker in GC. In one of the studies conducted by Ye et al, they used TCGA-STAD data that comprised of 370 samples. The results showed that the expression level of *LINC01094* is elevated in gastric cancer compared to normal tissues and found to be positively correlated with advanced stage. To determine the prognostic value of *LINC01094* expression, univariate and multivariate survival analyses were performed. The univariate analysis revealed a hazard ratio of 1.5 (95% CI: 1.06-2.05) with a *p*-value of 0.021, while the multivariate analysis resulted in a HR of 1.5 (95% CI: 1.02-2.30) and a *p*-value of 0.039. These findings indicate that elevated expression of *LINC01094* is associated with poor OS (23). In another study, Qi et al obtained data from 200 GC samples from the GEO database and demonstrated a correlation between the expression of *LINC01094* and OS. The HR was calculated to be 1.32 (95% CI: 1.06-1.64), with a significance level of 0.013 according to univariate survival analysis (19).

The importance of *CCDC144NL-AS1* and *LINC01094* have been also reported in other type of studies. By utilizing TCGA-STAD data, Sheng et al indicated that higher expression of *CCDC144NL-AS1* correlates with shorter OS in GC. Silencing of *CCDC144NL-AS1* significantly suppressed the cell proliferation and migration of GC cells, and also enhanced the chemosensitivity of gastric cancer cells to 5-fluorouracil (62). In agreement with studies showing that higher expression of *CCDC144NL-AS1* is associated with shorter overall survival of patients with gastric cancer, the prognostic value of *CCDC144NL-AS1* has been furthermore implicated in several other types of cancers including non-small cell lung cancer (63), colorectal (64) and ovarian (65) cancers as well as renal cell carcinoma (66).

On the other hand, *LINC01094* expression has been shown to be correlated with metastasis, T and N stages as well as poor OS in GC. Besides, knockdown of *LINC01094* prevents cellular growth and migration in GC (67). In the same way, correlation of elevated levels of *LINC01094* with poor survival has been reported in various cancer types like ovarian (68), pancreatic (69), breast (70) and colorectal (71) cancer.

This meta-analysis had several strengths. Firstly, most of the included studies had large sample size as well as long follow-up time. Secondly, most of the evaluated lncRNAs had other important features in their reported studies like being differential-expressed, involving in cancer-related pathways or having high degree centralities which further supports their role in cancer pathogenesis. Thirdly, there was a variation among the studies in the origin of samples (including TCGA, GEO, KM plotter, UALCAN, clinical samples, …) which can potentially provide better evidence of the validity of the obtained results in our meta-analysis. However, there were also some limitations: 1) A large number of included studies did not have complete information (samples size, *p*-value, hazard ratio or confidence interval) even after sending emails and so subsequently could not be entered to meta-analysis. 2) Few studies had performed multivariate analysis which considers cofounding factors and makes the results more precise and reliable. 3) Although origin of samples generally varied between different studies, some of the included studies, had utilized common data or databases such as TCGA or KM plotter which supposed to lead to similar results among these studies. Of note, compared to these studies, number of researches with analyzing survival rate based on their own collected clinical samples were limited.

## 5. Conclusions

In summary, the current study was the first systematic review and meta-analysis regarding the lncRNAs which have been introduced as potential prognostic biomarkers in GC based on ceRNA network studies. Many ceRNA network studies have revealed that dysregulation of lncRNAs expression may predict the survival rate of GC patients. In the current study, we systematically reviewed them and indicated this prognostic effect using meta-analysis. Furthermore, this meta-analysis indicated prognostic value of *CCDC144NL-AS1* and *LINC01094* in GC. Correlation of both of these lncRNAs with overall survival has been documented in other cancer types and seems that *CCDC144NL-AS1* and *LINC01094* are promising prognostic lncRNAs in the field of cancer. Further studies are required to validate the biological functions of these lncRNAs as prognostic biomarkers.

## Data Availability

This study is a systematic review and meta-analysis which conducted meta-analysis on published papers reporting ceRNA networks mainly based on the data of TCGA (https://portal.gdc.cancer.gov/) and GEO (https://www.ncbi.nlm.nih.gov/geo/) databases.

https://www.ncbi.nlm.nih.gov/geo/

## Acknowledgements

The Isfahan University of Medical Sciences in Isfahan, Iran, provided funding for this study.

## Ethical Approval and Consent to participate

This study is a systematic review and meta-analysis which conducted meta-analysis on published papers reporting ceRNA networks mainly based on the data of TCGA (https://portal.gdc.cancer.gov/) and GEO (https://www.ncbi.nlm.nih.gov/geo/) databases. The study protocol was approved by the Ethics Committee of Isfahan University of Medical Sciences and is available from here: https://ethics.research.ac.ir/ProposalCertificateEn.php?id=262634&Print=true&NoPrintHeader=true&NoPrintFooter=true&NoPrintPageBorder=true&LetterPrint=true.

## Consent for publication

Not applicable.

## Competing interests

The authors have no competing of interests to declare.

## Funding statement

The Isfahan University of Medical Sciences in Isfahan, Iran, provided funding for this study [grant number 140166].

## CRediT authorship contribution statement

**Sadra Salehi-Mazandarani:** Conceptualization, Investigation, Writing original draft, Writing-review & editing. **Mohammad Hossein Donyavi:** Investigation, Writing original draft, Writing-review & editing. **Amirhossein Vedaei:** Investigation, Writing original draft. **Alireza Najimi:** Formal analysis, Methodology, Writing original draft. **Ziba Farajzadegan:** Formal analysis, Methodology, Writing original draft. **Parvaneh Nikpour:** Supervision, Project administration, Writing original draft.

## Notes

### Competing Interest Statement

The authors have declared no competing interest.

