## Supplementary material for "Competing endogenous RNA networks reveal long non-coding RNAs as potential prognostic biomarkers in gastric cancer: A systematic review and meta-analysis": Suppl. File 1

| Supplementary table 1. Search strategy and number of records retrieved from each database | | |
| --- | --- | --- |
| Database | **Search query** | **Number of retrieved records up to 27 June 2022** |
| PubMed, all fields | (((cerna* OR "competing endogenous RNA" OR "competing endogenous RNAs" OR "competitive endogenous RNA" OR "competitive endogenous RNAs" OR "microRNA response element" OR "microRNA response elements") AND (network OR crosstalk)) OR ((lncrna* OR "long noncoding" OR long-noncoding* OR "long non-coding") AND (mirna* OR microRNA* OR micro-RNA*) AND (mrna* OR messenger-RNA*))) AND (Gastric OR stomach) AND (cancer OR carcinoma OR adenocarcinoma OR malignancy OR neoplasm OR tumor OR tumour) | 261 |
| Web of Science, topic | TS=((((cerna* OR competing-endogenous-RNA* OR competitive-endogenous-RNA* OR microRNA-response-element*) AND (network OR crosstalk)) OR ((lncrna* OR long-noncoding* OR long-non-coding*) AND (mirna* OR microRNA* OR micro-RNA*) AND (mrna* OR messenger-RNA*))) AND (Gastric OR stomach) AND (cancer OR carcinoma OR adenocarcinoma OR malignancy OR neoplasm OR tumor OR tumour)) | 491 |
| Scopus, title abstract keyword | TITLE-ABS-KEY((((cerna* OR "competing endogenous RNA*" OR "competitive endogenous RNA*" OR "microRNA response element*") AND (network OR crosstalk)) OR ((lncrna* OR long-noncoding* OR "long non-coding*") AND (mirna* OR microRNA* OR micro-RNA*) AND (mrna* OR messenger-RNA*))) AND (Gastric OR stomach) AND (cancer OR carcinoma OR adenocarcinoma OR malignancy OR neoplasm OR tumor OR tumour)) | 391 |
| Embase, all fields | (((cerna* OR competing-endogenous-RNA* OR competitive-endogenous-RNA* OR microRNA-response-element*) AND (network OR crosstalk)) OR ((lncrna* OR long-noncoding* OR long-non-coding*) AND (mirna* OR microRNA* OR micro-RNA*) AND (mrna* OR messenger-RNA*))) AND (Gastric OR stomach) AND (cancer OR carcinoma OR adenocarcinoma OR malignancy OR neoplasm OR tumor OR tumour) | 418 |
| Cochrane Library, title abstract | ((((cerna* OR "competing endogenous RNA*" OR "competitive endogenous RNA*" OR "microRNA response element*") AND (network OR crosstalk)) OR ((lncrna* OR long-noncoding* OR "long non-coding*") AND (mirna* OR microRNA* OR micro-RNA*) AND (mrna* OR messenger-RNA*))) AND (Gastric OR stomach) AND (cancer OR carcinoma OR adenocarcinoma OR malignancy OR neoplasm OR tumor OR tumour)):ti,ab,kw | 0 |
| ProQuest, title abstract | (((cerna* OR competing-endogenous-RNA* OR competitive-endogenous-RNA* OR microRNA-response-element*) AND (network OR crosstalk)) OR ((lncrna* OR long-noncoding* OR long-non-coding*) AND (mirna* OR microRNA* OR micro-RNA*) AND (mrna* OR messenger-RNA*))) AND (Gastric OR stomach) AND (cancer OR carcinoma OR adenocarcinoma OR malignancy OR neoplasm OR tumor OR tumour) | 96 |
| Google Scholar | ceRNA networks reveal lncRNAs as prognostic biomarkers in gastric cancer | First 100 hits |
