## Supplementary material for "Competing endogenous RNA networks reveal long non-coding RNAs as potential prognostic biomarkers in gastric cancer: A systematic review and meta-analysis": Suppl. File 2

**Supplementary table 2.** Results of quality assessment of 40 remaining studies^*^

| Ref | Q1 | Q2 | Q3 | Q4 | Q5 | Q6 | Q7 | Q8 | Q9 | Q10 | Q11 | Sum |
| --- | --- | --- | --- | --- | --- | --- | --- | --- | --- | --- | --- | --- |
| (1) | ✓ | ✓ | ✓ | ✓ | ✓ | ✓ | ✓ | ✓ | ✓ | ✓ | ✓ | 11 |
| (2) | ✓ | ✓ | ✓ | ✓ | ✓ | ✓ | ✓ | ✓ | 🗶 | ✓ | ✓ | 10 |
| (3) | ✓ | ✓ | ✓ | 🗶 | 🗶 | ✓ | ✓ | ✓ | 🗶 | ✓ | ✓ | 8 |
| (4) | ✓ | ✓ | ✓ | 🗶 | 🗶 | ✓ | ✓ | ✓ | ✓ | ✓ | ✓ | 9 |
| (5) | ✓ | ✓ | ✓ | ✓ | ✓ | ✓ | ✓ | ✓ | ✓ | ✓ | ✓ | 11 |
| (6) | ✓ | ✓ | ✓ | ✓ | ✓ | ✓ | ✓ | ✓ | ✓ | ✓ | ✓ | 11 |
| (7) | ✓ | ✓ | ✓ | 🗶 | 🗶 | ✓ | ✓ | ✓ | ✓ | ✓ | ✓ | 9 |
| (8) | ✓ | ✓ | ✓ | ✓ | ✓ | ✓ | ✓ | ✓ | ✓ | ✓ | ✓ | 11 |
| (9) | ✓ | ✓ | ✓ | ✓ | ✓ | ✓ | ✓ | ✓ | ✓ | ✓ | ✓ | 11 |
| (10) | ✓ | ✓ | ✓ | 🗶 | 🗶 | ✓ | ✓ | ✓ | 🗶 | 🗶 | 🗶 | 6 |
| (11) | ✓ | ✓ | ✓ | ✓ | 🗶 | ✓ | ✓ | ✓ | ✓ | ✓ | 🗶 | 9 |
| (12) | ✓ | ✓ | ✓ | ✓ | ✓ | ✓ | ✓ | ✓ | ✓ | ✓ | ✓ | 11 |
| (13) | ✓ | ✓ | ✓ | ✓ | ✓ | ✓ | ✓ | ✓ | 🗶 | ✓ | ✓ | 10 |
| (14) | ✓ | ✓ | ✓ | ✓ | 🗶 | ✓ | ✓ | ✓ | ✓ | ✓ | ✓ | 10 |
| (15) | ✓ | ✓ | ✓ | ✓ | ✓ | ✓ | ✓ | ✓ | 🗶 | ✓ | ✓ | 10 |
| (16) | ✓ | ✓ | ✓ | 🗶 | 🗶 | ✓ | ✓ | ✓ | ✓ | ✓ | 🗶 | 8 |
| (17) | ✓ | ✓ | ✓ | 🗶 | 🗶 | ✓ | ✓ | ✓ | ✓ | ✓ | ✓ | 9 |
| (18) | ✓ | ✓ | ✓ | 🗶 | 🗶 | ✓ | ✓ | ✓ | 🗶 | ✓ | 🗶 | 7 |
| (19) | ✓ | ✓ | ✓ | ✓ | ✓ | ✓ | ✓ | ✓ | 🗶 | ✓ | ✓ | 10 |
| (20) | ✓ | ✓ | ✓ | ✓ | 🗶 | ✓ | ✓ | ✓ | ✓ | ✓ | ✓ | 10 |
| (21) | ✓ | ✓ | ✓ | ✓ | ✓ | ✓ | ✓ | ✓ | ✓ | ✓ | 🗶 | 10 |
| (22) | ✓ | ✓ | ✓ | ✓ | 🗶 | ✓ | ✓ | ✓ | 🗶 | ✓ | 🗶 | 8 |
| (23) | ✓ | ✓ | ✓ | ✓ | ✓ | ✓ | ✓ | ✓ | ✓ | ✓ | ✓ | 11 |
| (24) | ✓ | ✓ | ✓ | 🗶 | 🗶 | ✓ | ✓ | ✓ | ✓ | ✓ | ✓ | 9 |
| (25) | ✓ | ✓ | ✓ | ✓ | ✓ | ✓ | ✓ | ✓ | ✓ | 🗶 | ✓ | 10 |
| (26) | ✓ | ✓ | ✓ | ✓ | 🗶 | ✓ | ✓ | ✓ | ✓ | ✓ | 🗶 | 9 |
| (27) | ✓ | ✓ | ✓ | 🗶 | 🗶 | ✓ | ✓ | ✓ | ✓ | ✓ | ✓ | 9 |
| (28) | ✓ | ✓ | ✓ | 🗶 | 🗶 | ✓ | ✓ | ✓ | 🗶 | ✓ | ✓ | 8 |
| (29) | ✓ | ✓ | ✓ | ✓ | ✓ | ✓ | ✓ | ✓ | ✓ | 🗶 | 🗶 | 9 |
| (30) | ✓ | ✓ | ✓ | 🗶 | 🗶 | ✓ | ✓ | ✓ | 🗶 | ✓ | 🗶 | 7 |
| (31) | ✓ | ✓ | ✓ | 🗶 | 🗶 | ✓ | ✓ | ✓ | ✓ | ✓ | ✓ | 9 |
| (32) | ✓ | ✓ | ✓ | ✓ | 🗶 | ✓ | ✓ | ✓ | 🗶 | ✓ | 🗶 | 8 |
| (33) | ✓ | ✓ | ✓ | 🗶 | 🗶 | ✓ | ✓ | ✓ | ✓ | ✓ | 🗶 | 8 |
| (34) | ✓ | ✓ | ✓ | ✓ | ✓ | ✓ | ✓ | ✓ | ✓ | ✓ | ✓ | 11 |
| (35) | ✓ | ✓ | ✓ | 🗶 | 🗶 | ✓ | ✓ | ✓ | ✓ | ✓ | ✓ | 9 |
| (36) | ✓ | ✓ | ✓ | 🗶 | 🗶 | ✓ | 🗶 | ✓ | ✓ | 🗶 | ✓ | 7 |
| (37) | ✓ | ✓ | ✓ | 🗶 | 🗶 | ✓ | ✓ | ✓ | 🗶 | 🗶 | ✓ | 7 |
| (38) | ✓ | ✓ | ✓ | 🗶 | 🗶 | ✓ | ✓ | ✓ | ✓ | ✓ | ✓ | 9 |
| (39) | ✓ | ✓ | ✓ | 🗶 | 🗶 | ✓ | ✓ | ✓ | ✓ | ✓ | ✓ | 9 |
| (40) | 🗸 | 🗸 | 🗸 | 🗸 | 🗸 | 🗸 | 🗸 | 🗸 | 🗶 | 🗸 | 🗸 | 10 |

^*^ Those studies (# 5) which did not pass this step, due to having low quality (quality score ≤7), have been red-highlighted.
