## Supplementary material for "Competing endogenous RNA networks reveal long non-coding RNAs as potential prognostic biomarkers in gastric cancer: A systematic review and meta-analysis": Suppl File 3

**Supplementary table 3.** Excluded articles based on the full text examination and quality assessment

| Reason | References |
| --- | --- |
| Low quality studies | (1-5) |
| Retracted studies | (6) |
| Other reasons |  |
| Specific conditions | (7-9) |
| Not English | (10, 11) |
| Full text not available | (12, 13) |
| Poster/Oral presentation | (14-16) |
| No survival analysis (or for lncRNAs) | (17-24) |
| No ceRNA network (or with lncRNA) | (25, 26) |
