## Supplementary material for "Competing endogenous RNA networks reveal long non-coding RNAs as potential prognostic biomarkers in gastric cancer: A systematic review and meta-analysis": Suppl File 4

| Supplementary table 4. The main characteristics of the 35 included studies | | | | | | | | | | |
| --- | --- | --- | --- | --- | --- | --- | --- | --- | --- | --- |
| First author | **Year** | **Country** | **Origin of samples** | **Sample size** | **lncRNA(s)** | **HR** | ***P*-value** | **CI** | **Follow-up**  **^(months)^** | **Ref.** |
| Ye | 2022 | China | TCGA | 370 (185, 185) | *LINC01094* | 1.476 | 0.021 | 1.060-2.054 | 125 | (1) |
| Huang | 2022 | China | TCGA | - | *CCDC144NL-AS1*  *HOXA10-AS*  *LINC01235* | 1.51  0.7  1.72 | 0.014  0.029  0.001 | 1.09-2.09  0.5-0.96  1.24-2.38 | 120 | (2) |
| Huangfu | 2022 | China | Clinical samples | 107 (53, 54) | *LINC00205* | 2.508 | 0.002 | 1.385-4.541 | 96 | (3) |
| Tang | 2022 | China | TCGA | 336 (168, 168) | *NKX2-1-AS1*  *VCAN-AS1*  *AC010145.1*  *LINC00469*  *C15orf54*  *IGF2-AS*  *AC018781.1*  *LINC00052*  *AL391152.1*  *POU6F2-AS2*  *ADAMTS9-AS1*  *ADAMTS9-AS2*  *FLRT1* | 0.997  1.065  0.996  0.993  1.003  0.999  1.006  1.002  1.010  1.003  1.003  1.0003  0.996 | 6.08e^-03^  1.02e^-02^  1.38e^-02^  1.68e^-02^  1.97e^-02^  2.14e^-02^  2.88e^-02^  2.94e^-02^  3.21e^-02^  3.75e^-02^  1.37e^-02^  1.97e^-02^  2.43e^-02^ | 0.990-1.004  1.035-1.095  0.979-1.012  0.974-1.014  0.997-1.008  0.996-1.002  1.0007-1.011  0.978-1.026  1.002-1.019  0.998-1.009  1.0007-1.005  0.999-1.002  0.979-1.014 | 120 | (4) |
| Ji | 2022 | China | GEPIA | 357 (184, 173) | *LINC00922* | 1.6 | 0.0056 | - | 120 | (5) |
| Piao | 2022 | China | Clinical samples | 50 (22, 28) | *HYPAL* | - | 0.00 | - | 100 | (6) |
| Liao | 2021 | China | TCGA | 375 | *AC010145.1*  *AC018781.1*  *ADAMTS9-AS1*  *ADAMTS9-AS2*  *AL139002.1*  *AL391152.1*  *IGF2-AS*  *LINC00326*  *POU6F2-AS2*  *VCAN-AS1*  *FRMD6-AS2*  *C15orf54* | -  1.145829  1.206821  1.119687  1.120617  1.167405  -  1.124694  1.0835  1.227419  1.145796  1.155017 | 4.085e^-2^  0.006166  0.001118  0.026685  0.002478  0.000913  1.186e^-2^  0.018552  0.023381  0.00132  0.013698  0.014314 | - | 120 | (7) |
| Wang | 2021 | China | TCGA  GEPIA  UALCAN | 343  GEPIA (379 (192, 187)  (100, 292) | *SNHG14*  *KCNQ1OT1*  *MUC19*  *ADAMTS9-AS2*  *ADAMTS9-AS2*  *PVT1*  *PVT1*  *PCBP1*  *LINC00461*  [*AC010336.1*](https://www.ncbi.nlm.nih.gov/nuccore/AC010336.1)  *AGAP11*  *HOTTIP*  *TTTY10*  *PCAT1* | -  1.5 | 0.068  0.367  0.209  0.003 (TCGA)  0.015 (GEPIA)  0.004 (TCGA)  0.018 (UALCAN)  0.605  0.175  0.165  0.392  0.249  0.130  0.553 | - | 120 (TCGA)  117 (GEPIA, UALCAN) | (8) |
| Guo | 2021 | China | TCGA | 379 | *H19*  *LINC00473*  *AC079160.1*  *AC093866.1* | - | 0.36577  0.09563  0.05342  0.07788 | - | 120 | (9) |
| Ye | 2021 | China | TCGA | 378 | *LINC02731*  *MIR99AHG*  *INHBA-AS1*  *LINC02613*  *CCDC144NL-AS1*  *VLDLR-AS1*  *LINC01497*  *LIFR-AS1*  *A2M-AS1*  *LINC01537*  *LINC00702* | 1.527  1.447  1.419  1.480  1.439  1.379  1.596  1.466  1.472  1.502  1.457 | 0.036  0.008  0.024  0.023  0.018  0.030  0.045  0.005  0.044  0.002  0.025 | 1.109-2.103  1.052-1.992  1.034-1.949  1.078-2.032  1.046-1.981  1.004-1.896  1.159-2.198  1.066-2.016  1.070-2.024  1.093-2.064  1.057-2.007 | 133 | (10) |
| Zhang | 2021 | China | TCGA | 345 | *HMGA2-AS1*  *LINC01446*  *LINC01644*  *C7orf65*  *LINC01537*  *LINC01981*  *CASC20*  *LOC105373764*  *ABCA9-AS1*  *LOC105375787*  *LINC02182*  *LINC02266*  *LINC01697*  *LOC339260*  *CCDC144NL.AS1*  *CYMP.AS1*  *LOC101929532*  *LINC02042*  *HOXA11.AS*  *ADAMTS9-AS1*  *LINC02657*  *LINC02389*  *LOC100506388*  *LOC105369201*  *LINC02268*  *FLJ42969*  *TMEM132D-AS1*  *LOC101928924*  *LINC01060*  *LINC02465*  *LINC02269*  *LINC01606*  *LINC01592*  *LINC01146*  *LINC01235* | - | 0.0315  0.0034  0.0490  0.0092  0.0087  0.0253  0.0376  0.0476  0.0417  0.0426  0.0352  0.0199  0.0164  0.0067  0.0067  0.0187  0.0308  0.0167  0.0432  0.0093  0.0402  0.0408  0.0246  0.0252  0.0387  0.0195  0.0157  0.0331  0.0447  0.0103  0.0212  0.0185  0.0214  0.0451  0.0130 | - | 100 | (11) |
| Teng | 2021 | China | Clinical specimens | 178 (89, 89) | *NKX2‐1‐AS1* | 0.358 | **<0.001** | 0.261–0.448 | 100 | (12) |
| Fan | 2020 | China | KM plotter | - | *CCDC144NL-AS1* | 1.59 | **0.0035** | 1.23-2.06 | 150 | (13) |
| Peng | 2020 | China | TCGA | 361 | *PVT1*  *ZNF667-AS1*  *HAND2-AS1* | - | -  -  0.046 | - | 117 | (14) |
| Sun | 2020 | China | TCGA | 180 | *LINC00330*  *AC061975.6*  *AP002478.1*  *ST7-AS2*  *AC123777.1*  *LINC00346*  *LINC00473*  *AC007389.1*  *AL158206.1*  *LINC00365*  *PVT1*  *TM4SF19-AS1*  *AC110491.1*  *DSCR4-IT1*  *LINC00460*  *AC011374.1*  *HCG22* | 1.117  1.067  1.070  1.096  1.822  1.019  1.047  1.573  1.005  1.033  0.990  1.072  1.363  1.333  1.020  1.202  1.009 | 0.000  0.000  0.003  0.004  0.005  0.007  0.017  0.021  0.022  0.024  0.028  0.031  0.042  0.043  0.044  0.045  0.049 | 1.064-1.173  1.030-1.106  1.023-1.119  1.030-1.165  1.196-2.778  1.005-1.033  1.008-1.086  1.070-2.311  1.001-1.009  1.004-1.062  0.981-0.999  1.006-1.142  1.011-1.837  1.009-1.761  1.001-1.040  1.004-1.439  1.000-1.018 | 84 | (15) |
| Zhang | 2020 | China | TCGA | - | *MAGI2-AS3*  *HMGA1P4*  *UBE2Q1-AS1*  *MIR22HG* | 1.387 | 0.047  0.043  0.909  0.527 | 1.005–1.914 | 120 | (16) |
| Mao | 2020 | China | TCGA  Clinical samples | 349  112 | *LINC01106*  *FOXD2-AS1*  *AC103702.2*  *LINC01106*  *FOXD2-AS1* | - | 0.0061  0.03  0.0068  0.0091  0.0023 | - | 133(TCGA)  50(Clinical specimens) | (17) |
| Liu | 2020 | China | TCGA | RNA (343) | *ADAMTS9-AS1*  *C15orf54*  *AL391152.1* | - | 6.721e^-03^  2.041e^-02^  2.045e^-02^ | - | 120 | (18) |
| Li | 2020 | China | TCGA, UALCAN | 392 (104, 288) | *H19* | - | 0.011 | - | 100 | (19) |
| Zu | 2020 | China | KM plotter | DLGAP1-AS1 (631 (237, 394)  PVT1 (631 (294, 337)  RECQL4 (876 (247, 629))  HCG18 (631 (411, 220)) | DLGAP1-AS1  PVT1  RECQL4  HCG18 | 1.77  1.67  1.28  2.07 | 1.6e^-07^  2.6e^-06^  0.0093  9.6e^-09^ | 1.43-2.2  1.34-2.07  1.06-1.54  1.61-2.67 | 150 | (20) |
| Qi | 2020 | China | TCGA | 372 | *AC018647.3*  *MAGI2‐AS3*  *MIR99AHG*  *NR2F1‐AS1*  *LINC00106*  *PVT1*  *RP5‐1074L1.4*  *RP11‐7K24.3* | - | 0.00049  0.015  0.014  0.014  0.013  0.015  0.033  0.035 | - | 133 | (21) |
| Nie | 2020 | China | KM plotter | HCG18 (631 (411, 220))  SNHG16 (631 (296, 335))  H19 (631 (378, 253))  SNHG3 (876 (393, 483))  HNRNPU-AS1 (631 (322, 309)) | *HCG18*  *SNHG16*  *H19*  *SNHG3*  *HNRNPU-AS1* | 2.07  1.45  1.96  2.05  1.37 | 9.6e-09  0.00065  1.2e-08  <1e-16  0.0044 | 1.61-2.67  1.17-1.8  1.55-2.48  1.73-2.44  1.1-1.71 | 150 | (22) |
| Shen | 2020 | China | UALCAN  Clinical specimens | UALCAN (392 (104, 288))  50 (25, 25) | *H19* | - | 0.011  0.018 | - | 100(UALCAN)  80(Clinical specimens) | (23) |
| Zhang | 2020 | China | TCGA | - | *MIR4458HG*  *LINC01235* | - | 0.21  0.008 | - | 133 | (24) |
| Qi | 2019 | China | GEO (GSE15459) | 193 | *LL22NC03-N14H11.1*  *RP3-428L16.2*  *LINC01105*  *LINC00982*  *LINC01094*  *MIR100HG*  *DNM3OS*  *GATA6-AS1*  *RP11-363E7.4* | 1.29  1.2  0.7  0.74  1.32  1.15  1.2  0.85  0.86 | < 0.001  0.004  0.010  0.011  0.013  0.014  0.020  0.028  0.032 | 1.11-1.50  1.06-1.35  0.54-0.92  0.59-0.93  1.06-1.64  1.03-1.28  1.03-1.41  0.73-0.98  0.75-0.99 | 150 | (25) |
| Peng | 2019 | China | TCGA | 365 | *MIR100HG*  *MAGI2-AS3*  *AC080038.1*  *AC010478.1*  *MEF2C-AS1*  *RAP2C-AS1*  *AC022034.2*  *AC104825.2*  *NR2F1-AS1*  *MBNL1-AS1*  *AC135012.3*  *ADAMTS9-AS2*  *PART1*  *SNHG14*  *PWAR6*  *AC067750.1*  *CTD-2201I18.1*  *MIR99AHG*  *AC005358.2*  *HAND2-AS1*  *LINC01266*  *LINC01354*  *AC008808.2*  *AC079789.1*  *AC105942.1*  *AC124312.5*  *AC233702.10*  *GAS1RR*  *LYPLAL1-AS1*  *ZNF667-AS1*  *AC007495.1*  *AL691447.2*  *FENDRR*  *RBMS3-AS3*  *AC006059.1*  *AC016722.3*  *AC025165.1*  *AC120049.1*  *AL139147.1*  *AP001528.2*  *CADM3-AS1*  *A2M-AS1*  *AC079630.1*  *AL356599.1* | 1.048411  1.242736  3.360686  1.182393  1.749401  1.050688  0.969829  1.056401  1.288318  0.991053  1.245253  1.042312  1.063839  1.166315  0.997848  1.047768  1.319286  1.617249  6.525081  0.984551  3.270522  2.658016  1.040367  1.566297  1.021449  1.0101  3289.664  2.448866  0.715678  1.038709  1.699957  1.542672  1.011148  1.083634  1.419768  1.097406  1.232356  1.392001  5.839658  1.547054  2.802363  1.011894  2.810959  0.679064 | 0.175386  0.06959  0.065983  0.385268  0.552753  0.822708  0.614905  0.573507  0.159086  0.737805  0.629784  0.899259  0.620626  0.407183  0.989369  0.901524  0.336403  0.037948  0.257969  0.605558  0.342831  0.228987  0.728891  0.392122  0.650374  0.948257  0.287533  0.227506  0.774458  0.601017  0.577916  0.40415  0.650783  0.864213  0.335101  0.845642  0.676442  0.242593  0.016491  0.05401  0.064674  0.929037  0.173474  0.504824 | - | 117 | (26) |
| Pan | 2019 | China | TCGA | 368  *ADAMTS9-AS2* (112,256)  *ARHGEF26-AS1* (115,253)  *HOTAIR* (215,153)  *HOTTIP* (207,161)  *LINC00052* (237,131)  *NKX2-1-AS1* (180,188)  *VCAN-AS1* (127,241) | *ADAMTS9-AS2*  *ARHGEF26-AS1*  *HOTAIR*  *HOTTIP*  *LINC00052*  *NKX2-1-AS1*  *VCAN-AS1* | 0.55  1.45  1.6  0.7  1.43  1.46  1.88 | 0.003  0.034  0.006  0.033  0.046  0.022  <0.001 | 0.39-0.78  1-2.09  1.15-2.22  0.51-0.98  1.02-2  1.05-2.03  1.32-2.68 | 133 | (27) |
| Zhang | 2019 | China | TCGA | 367 (183, 184) | *HMGA1P4* | - | 0.026 | - | 120 | (28) |
| Yang | 2019 | China | GEPIA | RP11-169F17.1 (355 (190, 165))  RP11-669N7.2 (366 (190, 176) | *LINC01234*  *HOXC-AS3*  *RP11-400N13.2*  *RP11-138J23.1*  *HOTAIR*  *RP11-191L9.4*  *AC007128.1*  *RP11-476K15.1*  *LINC01050*  *NPSR1-AS1*  *CTC-480C2.1*  *KCNMB2-AS1*  *HOXC-AS2*  *CTD-2147F2.1*  *RP11-366F6.2*  *RP4-594A5.1*  *RP11-742B18.1*  *LINC01614*  *LINC00355*  *RP11-254F7.1*  *RP11-297P16.4*  *RP11-89K21.1*  *HCG22*  *RP11-169F17.1*  *RP11-197K6.1*  *RP11-474D1.3*  *AC007099.1*  *AF127577.8*  *RP6-114E22.1*  *FEZF1-AS1*  *RP5-884M6.1*  *RP11-962G15.1*  *RP11-346D19.1*  *RP11-669N7.2*  *LINC01194*  *AC012363.4*  *AC079466.1*  *LL22NC03-N64E9.1*  *CTD-2591A6.2*  *CTD-2532K18.2*  *AC012531.25*  *RP11-874J12.4*  *RP11-115D19.1*  *RP11-867G2.6*  *LINC00858*  *AC068580.7*  *LINC01163*  *RP11-116D2.1*  *CTD-2331C18.5*  *RP11-190J1.3*  *LHX5-AS1*  *AC079630.2* | 1.6  1.4 | 0.28  0.76  0.055  0.99  0.099  0.96  0.27  -  0.15  0.18  0.14  0.21  0.63  0.045  -  0.12  0.32  0.02  -  0.51  -  0.81  0.28  0.0066  -  0.36  0.81  0.18  0.079  0.94  0.12  -  0.83  0.032  -  0.15  0.36  0.86  -  -  0.94  0.99  0.17  0.29  0.6  0.15  0.34  -  -  0.99  -  - | - | 120 | (29) |
| Zhu | 2019 | China | TCGA | - | *SNHG4*  *LINC00523* | 1.54  1.6 | 8.9e-05  2.2e-05 | 1.24-1.91  1.29-2 | 150 | (30) |
| Wang | 2019 | China | GEPIA  TCGA | 379 (192, 187)  373 | *ADAMTS9-AS2* | 1.5  - | 0.014  4.756e^-03^ | -  - | 120  120 | (31) |
| Arun | 2018 | India | TANRIC | 226 | *HOTAIR*  *GAS5*  *LINC00312*  *CCAT1*  *ZEB2-AS1*  *TUG1*  *H19*  *MEG3*  *LINCROR*  *MALAT1*  *NEAT1*  *UCA1* | - | 0.0380  0.4187  0.5632  0.8358  0.8985  0.4229  0.8494  0.8222  0.8090  0.9699  0.7438  0.6247 | - | - | (32) |
| Wang | 2018 | China | KM plotter | 631 (221, 410) | *RP11-363E7.4* | 0.69 | 0.0019 | 0.55-0.88 | 150 | (33) |
| He | 2018 | China | TCGA | 372 | *VCAN‑AS1*  *IGF2‑AS*  *ABCA9‑AS1*  *ADAMTS9‑AS1*  *ADAMTS9‑AS2*  *AL391152.1*  *AL139002.1*  *C15orf54*  *FRMD6‑AS2* | - | 0.005  0.009  0.010  0.014  0.019  0.024  0.031  0.042  0.043 | - | 120 | (34) |
| Li | 2016 | China | TCGA | 280 | *RPLP0P2*  *FOXD2-AS1*  *H19*  *TINCR*  *SLC26A4-AS1*  *SMIM10L2A*  *SMIM10L2B*  *SNORD116-4* | - | 0.0001  0.022  0.002  0.016  0.007  0.029  0.001  0.037 | - | 67 | (35) |

*GEO: Gene Expression Omnibus; GEPIA: Gene Expression Profiling Interactive Analysis; KM plotter: Kaplan Meier plotter; lncRNA: long non-coding RNA; TANRIC: The Atlas of Noncoding RNAs in Cancer; TCGA: The Cancer Genome Atlas; UALCAN: University of ALabama at Birmingham CANcer data analysis Portal
